# Multi-omics and network propagation reveal latent innate immune programmes stratifying high-risk thrombotic primary antiphospholipid syndrome

**DOI:** 10.64898/2026.06.26.26356680

**Authors:** Srijith Sasikumar, Michail Baltsiotis, Kleio-Maria Verrou, Georgia Rouni, Petros P. Sfikakis, Martina Samiotaki, Evangelia Petsalaki, Maria G. Tektonidou

## Abstract

**Objectives:** Thrombotic primary antiphospholipid syndrome (thrPAPS) outcomes are associated with thrombosis type (arterial versus venous), recurrence, and antiphospholipid antibody (aPL) profile (single versus triple-aPL). We investigated molecular signatures underlying disease status and high-risk phenotypes.

**Methods:** We performed whole-blood transcriptomics and mass spectrometry–based plasma proteomics in patients with thrPAPS and age/sex-matched healthy controls. Analyses included differential expression, pathway enrichment, weighted gene co-expression network analysis (WGCNA) and machine learning. Multi-Omics Factor Analysis (MOFA2) and network propagation were applied to identify latent molecular programmes associated with high-risk phenotypes.

**Results:** Transcriptomic and WGCNA analyses revealed an interferon-associated module associated with high-risk phenotypes. Plasma proteomics distinguished thrPAPS from healthy controls through a coordinated thromboinflammatory signature encompassing complement, acute-phase, platelet, and coagulation-associated pathways. Complement factor D, a rate-limiting enzyme of the alternative complement pathway, discriminated recurrent from single-event thrPAPS (AUC = 0.79) and correlated with thrombotic event count (Spearman’s ρ = 0.62, p < 0.001). Mixed arterial/venous phenotype showed the greatest degree of subgroup-specific dysregulation, including complement and coagulation/fibrinolysis-related proteins. MOFA2 identified a proteome-dominant latent factor that increased with aPL burden (Spearman’s ρ = 0.33, p = 0.017) and was enriched for complement cascade proteins. Network propagation embedded this signature within immune-cell signalling (STAT-1, PI3K–AKT, MAPK8, SRC), N-linked glycosylation, and mitochondrial oxidative phosphorylation. Longitudinal profiling identified reactive oxygen species–associated proteins during active disease.

**Conclusions:** ThrPAPS is characterised by a complement-, interferon and platelet-driven thromboinflammatory programme that scales with aPL and thrombosis burden, converging on innate immune activation as a central feature of high-risk disease.

**WHAT IS ALREADY KNOWN ON THIS TOPIC:**

- Thrombotic primary antiphospholipid syndrome (thrPAPS) is a rare and potentially fatal autoimmune disorder carrying a high risk of thrombotic recurrence, often despite adequate anticoagulant treatment, and an increased mortality risk. A few previous studies investigated transcriptomics or proteomics in antiphospholipid syndrome, but many included mixed populations of various antiphospholipid syndrome groups (obstetric, thrombotic, or microvascular APS and/or asymptomatic antiphospholipid antibody carriers) or both primary and secondary antiphospholipid syndrome.
- The molecular programmes linking circulating proteomic changes to cellular transcriptional states in patients with exclusively primary antiphospholipid syndrome and a history of thrombosis remain uncertain, as do the associations with high-risk phenotypes associated with poor outcomes.

**WHAT THIS STUDY ADDS:**

- This study integrates, for the first time, plasma proteomics with whole-blood transcriptomics to identify molecular determinants of high-risk phenotypes associated with worse outcomes. Plasma proteomics separated thrPAPS from healthy controls through a coordinated thromboinflammatory signature spanning complement, acute-phase, platelet, and coagulation-associated pathways, with this signature reproducing consistently across differential abundance, co-expression network, and latent-factor analyses.
- Complement factor D (CFD), the rate-limiting enzyme of the alternative complement pathway, emerged as a quantitative correlate of thrombotic burden (Spearman ρ = 0.62, p < 0.001) and discriminated recurrent from single-event disease (AUC = 0.79), nominating it as a mechanistically anchored candidate marker of recurrence risk for future validation.
- Transcriptomic and WGCNA analyses revealed an interferon-associated module associated with high-risk phenotypes. Multi-omics factor analysis identified a proteome-dominant latent programme that increased monotonically with aPL burden and was enriched for the complement cascade; network propagation embedded this programme within innate immune-receptor signalling (STAT1, SRC, PI3K–AKT, MAPK8), linking circulating signatures to interferon-associated transcriptional states identified within the same cohort.
- Longitudinal paired sampling of active versus non-active disease identified reactive oxygen species–associated proteins as features of disease activity

**HOW THIS STUDY MIGHT AFFECT RESEARCH, PRACTICE OR POLICY:**

- These findings provide a molecular framework for stratifying high-risk thrPAPS phenotypes. The identified proteomic and multi-omics integration markers and pathways may inform future biomarker validation studies and guide the development of personalised, targeted therapeutics.

## Introduction

Antiphospholipid syndrome (APS) is a systemic autoimmune disorder characterised by thrombotic, microvascular and obstetric manifestations in the presence of persistently positive antiphospholipid antibodies (aPL), including lupus anticoagulant, anticardiolipin antibodies, and anti-β2-glycoprotein I antibodies (1). Thrombotic APS is associated with substantial morbidity and mortality, with recurrent events often occurring despite anticoagulation treatment (2). Poor outcomes, including damage accrual and mortality in thrombotic APS, are independently associated with thrombosis type (arterial versus venous thrombosis), thrombotic event recurrence, and triple aPL positivity (3–6). Pathogenic aPL antibodies promote a proinflammatory and prothrombotic state by activating endothelial cells, monocytes, neutrophils, and platelets (7–10). However, the molecular signatures underlying thrombotic APS phenotypes at high risk of poor outcomes remain incompletely defined.

A few high-throughput transcriptomic studies have begun to define molecular endotypes of thrombotic APS (11). Primary APS provides an ideal model for such investigations, by minimising confounding from co-existing autoimmune disorders. We previously showed that whole-blood RNA sequencing in thrombotic primary APS (thrPAPS) identified type I and II interferon-regulated genes as key transcriptional drivers of the disease (12) and highlighted STAT-1 as a potential treatment target (13). Other studies have similarly implicated interferon signalling (14–16), while more recent kidney tissue (17) and whole-blood transcriptomic studies have highlighted neutrophil extracellular trap (NET) pathways (18). However, transcriptomics primarily captures cellular transcriptional states rather than downstream functional activity. Plasma proteomics provides complementary information by profiling circulating proteins released from activated cells and tissues. A recent plasma proteomic study revealed pathways related to innate and adaptive immune systems and extracellular matrix organization across aPL+/NonAPS, thrombotic APS, and microvascular APS subtypes (10 samples/group) (19). Multiomics integration can provide a comprehensive view of disease mechanisms by defining molecular programmes that link cellular activation states to circulating effector pathways.

Here, we integrated whole-blood transcriptomics and plasma proteomics with a network propagation approach in a well-characterised thrPAPS patient cohort to identify molecular programmes associated with disease status, high-risk phenotypes, and disease activity.

### Patients and Methods

We included 59 of 62 patients with thrombotic primary APS (thrPAPS) (1) who participated in our previous transcriptome studies (12,13) and had paired plasma samples for proteomic analysis. Twenty-nine age- and sex-matched healthy controls (HCs) were included. Plasma proteomic data were analysed for 55 of the 59 patients with thrPAPS and 23 age- and sex-matched healthy controls after excluding samples that failed quality control (**Supplementary Table 1**). Exclusion criteria were co-existing autoimmune rheumatic disorders, treatment with biologics or glucocorticoids, infections, malignancies, and pregnancy. Patient and public involvement was not included in this study.

### Preprocessing of whole-blood RNA-Sequencing data

Sample collection, RNA extraction, library preparation, and sequencing procedures have been reported previously (12). For the present study, raw FASTQ files from 59 patients and 29 HCs were re-analysed as described in the **Supplementary Methods**. Transcript abundance was quantified using Salmon (20), aggregated to gene-level counts, and analysed using DESeq2 (21). Variance-stabilising transformation (VST) was applied to the gene expression matrix for downstream analyses, including clustering, pathway enrichment, and integration with proteomic data. (**Supplementary Table 2**).

### Mass spectrometry-based plasma label-free proteomics

Plasma samples (50 μL) were processed using the ENRICHplus kit (PreOmics, Germany). Detailed sample preparation procedures are provided in the **Supplementary Methods**. Peptide samples were analysed by nanoLC-MS/MS using a nanoElute 2 liquid chromatography system coupled online to a timsTOF Ultra 2 mass spectrometer (Bruker Daltonik GmbH). Peptides were separated on a temperature-controlled (50 °C) Aurora C18 column (25 cm × 75 μm i.d., 1.7 μm; IonOpticks) with a 30-min gradient at 300 nL/min. The instrument operated in dia-PASEF mode using the low-input sample method (mobility range 0.64–1.45; 0.96 s cycle time). Full MS1 spectra were acquired across 100–1700 m/z, with ion mobility separation employing 3 isolation windows per TIMS scan (8 scans covering 24 total windows).

### Proteomics database searching

Raw timsTOF dia-PASEF data were processed in Bruker ProteoScape using the integrated Spectronaut 19.5 DIA workflow against the reviewed UniProt human proteome (20,578 entries; retrieved May 29, 2025). Default search settings were applied, permitting up to two missed tryptic cleavages. Cysteine carbamidomethylation was set as a fixed modification, while variable modifications included N-terminal protein acetylation, asparagine/glutamine deamidation, and methionine oxidation. The Q cut-off values for DIA identification, both at the precursor and protein level, were set to 0.01. Run-wise reprocessing was performed individually using a global library, enabling run-to-run matching (MBR).

### Preprocessing and imputation of missing data

A total of 2,187 proteins were quantified across all samples, with a minimum of 370 per sample. Technical replicates were available for 20 samples (a maximum of 2 runs per sample); these were collapsed to the median of log2-transformed intensities, and replicate concordance was assessed using Pearson correlation (**Supplementary Table 3)**. Proteins missing in more than 80% of samples were excluded (22). The remaining missing values were imputed using PIMMS (Proteo-omics Imputation using Self-Supervised deep learning), a method that leverages non-random missingness patterns common in label-free proteomics (23,24) **(Supplementary Table 4).**

### Differential protein expression analysis

Differential protein abundance was analysed using the limma package (25) with linear models, with disease status as the primary variable and sex and age as covariates. Statistical significance thresholds were set at adjusted p-value < 0.05 and |log2 fold change| > 1. Separate models were fitted for thrombosis location, the number of thrombotic events, and aPL burden. For the recurrent vs single-event thrombosis comparison, models were additionally adjusted for treatment, and significance was defined as LFC >0.5 and a nominal p-value <0.001, given the smaller group sizes. Pathway enrichment analysis was performed using Metascape (26) and STRING (27).

### Gene co-expression network analysis

Imputed proteomic data were integrated with sample metadata and filtered to retain samples and proteins that passed quality control in both the thrPAPS and HC groups. A weighted gene co-expression network (WGCNA) was constructed using the WGCNA package (28) on the combined dataset. A signed network was generated with a soft-thresholding power of 8, minimum module size of 30, and dynamic tree cutting (deepSplit = 3, mergeCutHeight = 0.15). Associations between module eigengenes and thrPAPS status (a binary trait) were assessed using Pearson’s correlation. P values were adjusted using the Benjamini–Hochberg method, and modules were ranked by significance and effect size. Transcriptomic WGCNA is described in the **Supplementary Methods**.

### Machine learning classification models

Classification models were built using features derived from WGCNA protein modules in Python 3. The methodology is presented in the **Supplementary Methods.**

### Multi-omics factor analysis

MOFA2 (29) was applied to integrate plasma proteomics and whole-blood transcriptomics (DESeq2 variance-stabilising transformation) data from 55 thrPAPS patients with complete data in both modalities. Before modelling, proteins with variance below the 30th percentile were excluded. For the transcriptomics view, canonical haemoglobin genes were removed to reduce contamination from erythroid cells, and the 5,000 most variable genes were selected. MOFA2 training is described in the **Supplementary Methods**.

### Network propagation using phuEGO and functional module analysis

The top 100 positively weighted proteins contributing to each factor were selected as seed nodes. Network propagation was performed using phuEGO (30) on a human protein–protein interaction network. Propagated subnetworks were partitioned into functional modules using the phuEGO framework. Functional enrichment of individual modules was assessed using over-representation analysis against Hallmark, Reactome and Gene Ontology Biological Process gene sets.

## Results

### An interferon transcriptional programme defines high-risk thrPAPS phenotypes

To establish the transcriptomic baseline and identify co-expression modules associated with high-risk phenotypes, we re-analysed whole-blood transcriptomic data from 59 patients and 29 age- and sex-matched healthy controls (all White) (12). Differential gene expression analysis (|log₂FC| > 1, adjusted p < 0.05) identified type I and II interferon-regulated genes (IRGs) as the predominant upregulated transcripts in thrPAPS, while MYOM2 was among the five downregulated genes, consistent with findings from our previous study (**Supplementary Table 5**).

In a WGCNA analysis within the thrPAPS cohort, we used variance-filtered whole-blood transcriptomic data and assessed module-trait associations with high-risk phenotypes. This analysis identified the pink module as significantly associated with triple-aPL (r = 0.38, p = 0.007) and inversely associated with single-aPL positivity (r = −0.32, p = 0.03) (Supplementary Figure 1A; Supplementary Table 6). Functional enrichment analysis demonstrated strong overrepresentation of interferon-related pathways in this module, including interferon signalling, the antiviral innate immune response, and antimicrobial mechanisms of interferon-stimulated genes (**Supplementary Figure 1B**).

### Plasma proteomics reveal a distinctive complement, acute phase response and coagulation-associated molecular signature in thrPAPS

In total, 2,187 proteins were quantified, with a minimum of 370 proteins detected per sample. After exclusion of proteins missing in more than 80% of samples, 1,447 proteins were retained for downstream analysis. The remaining missing values were imputed using the PIMMS framework before statistical analysis (23,24).

Principal component analysis showed clear separation between thrPAPS and HCs (**Figure 2A**). Differential abundance analysis using Limma, adjusted for age and sex (**Materials & Method**s), identified 148 upregulated and 302 downregulated proteins in thrPAPS relative to HCs (adjusted p<0.05, |log2FC|>1; **Figure 2B, C; Supplementary Table 7**). Upregulated proteins included multiple components linked to complement regulation, coagulation, and the acute-phase response, including SERPINA1, SERPINA3, SERPINC1, SERPING1 and RBP4. Pathway enrichment analysis showed that proteins increased in thrPAPS were strongly enriched for complement, platelet degranulation and acute-phase response pathways (**Figure 2D**).

**Figure 1.**
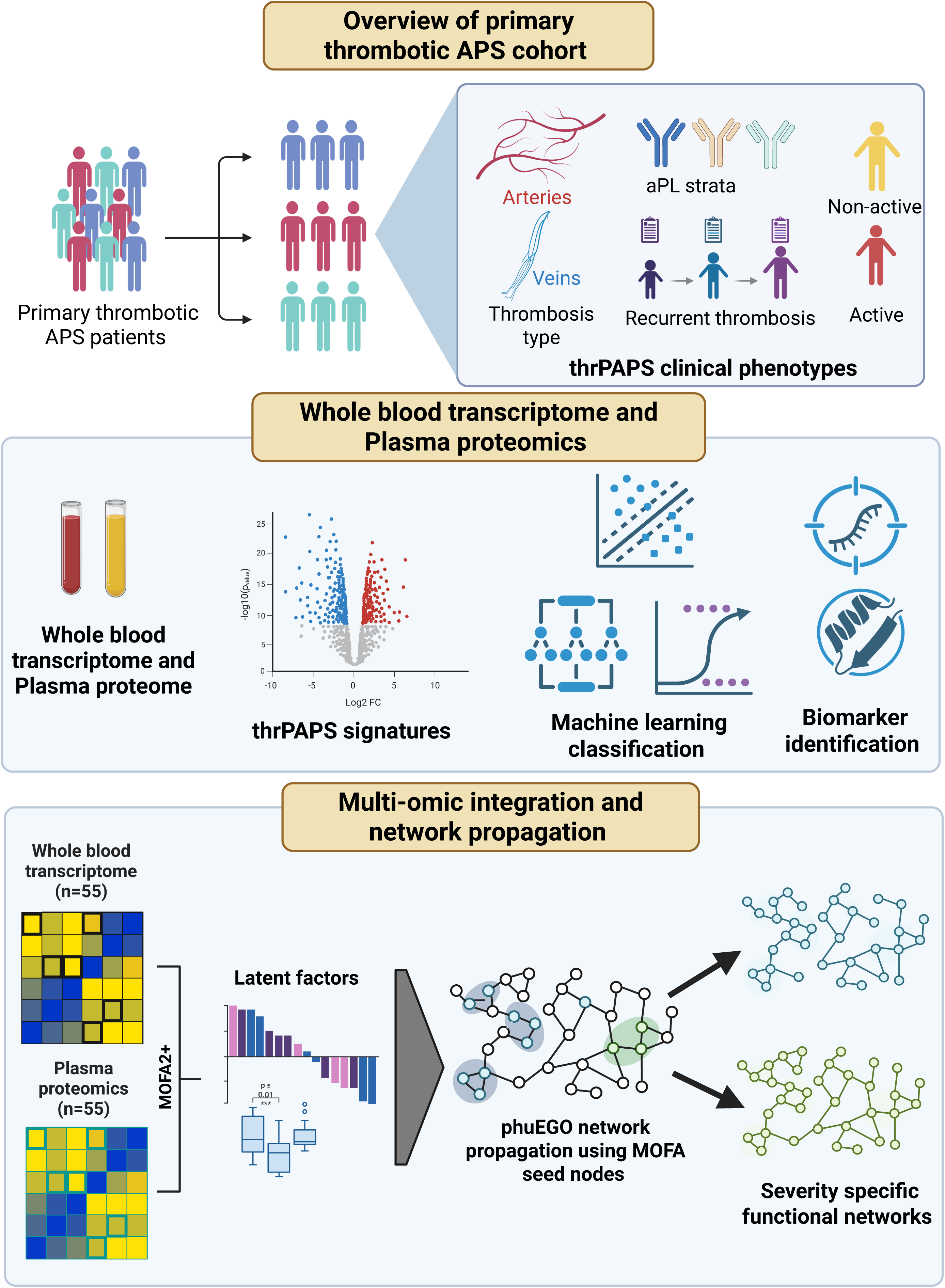
Overview of the multi-omic study design for characterising thrombotic primary antiphospholipid syndrome (thrPAPS). Patients with thrPAPS were stratified by clinical phenotype: thrombosis type (arterial, venous, or combined arterial–venous), antiphospholipid antibody (aPL) profile (single-, double-, or triple-positive), thrombotic recurrence (single versus recurrent events), and disease activity status (active versus non-active). Plasma proteomes and whole-blood transcriptomes from thrPAPS and healthy controls were profiled to define transcriptomic and plasma signatures, train machine-learning classifiers, and identify candidate biomarkers. Paired plasma proteomic and whole-blood transcriptomic data from 55 patients were integrated using Multi-Omics Factor Analysis (MOFA2) to identify latent molecular programmes, and phuEGO network propagation with MOFA-derived seed nodes was used to reconstruct severity-specific functional networks (Created with Biorender.com)

**Figure 2.**
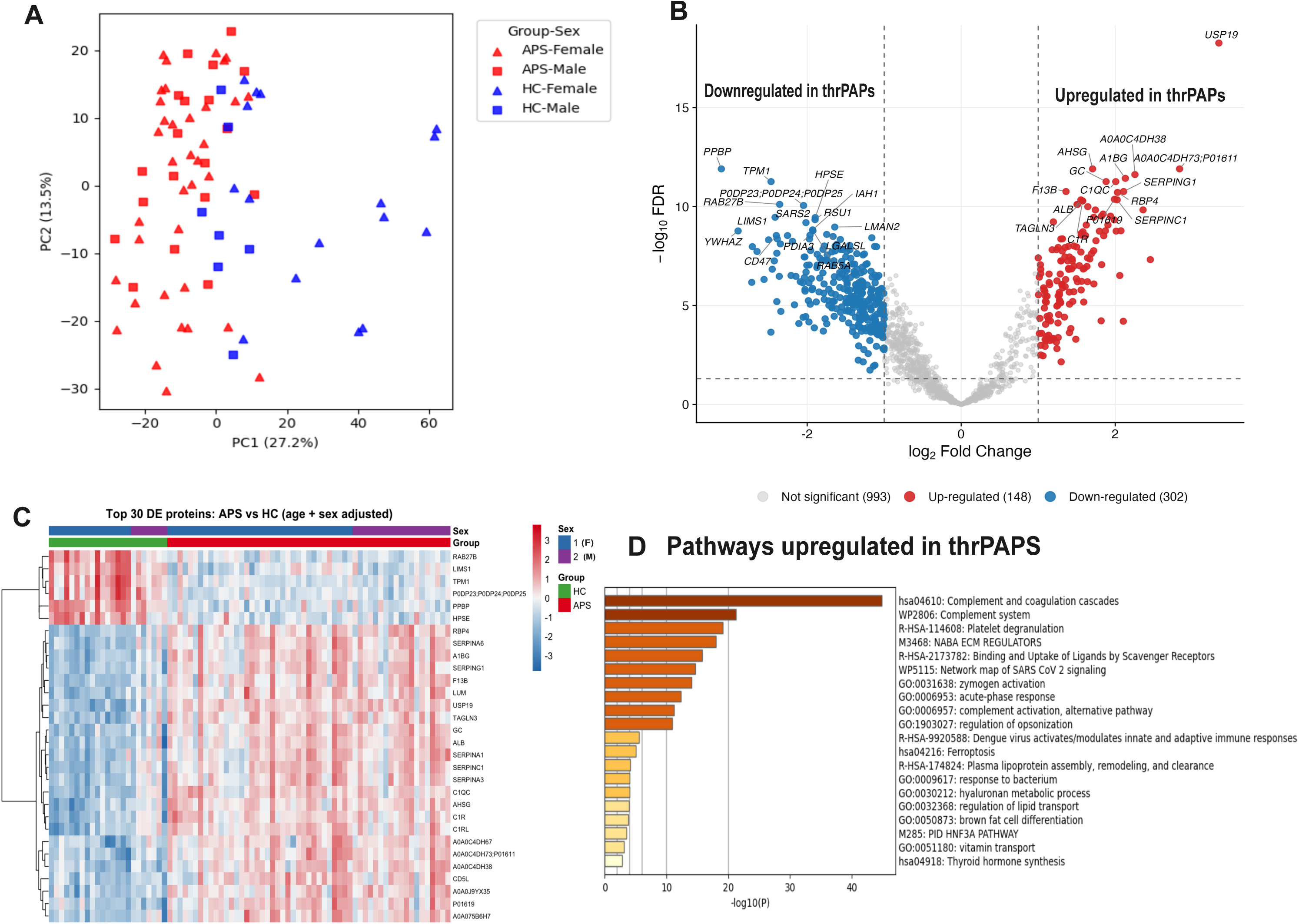
Plasma proteomic signatures of thrPAPS. (A) Principal component analysis (PCA) of plasma proteomes from thrPAPS patients (red) and healthy controls (HCs, blue), separated by sex (triangles = female; squares = male), showing clear separation along PC1. (B) Volcano plot of differential protein expression between thrPAPS patients and HCs (limma, age- and sex-adjusted; FDR ≤ 0.05, |log₂FC| ≥ 1). Red = upregulated (n=148); blue = downregulated (n=302); grey = not significant (n=993). (C) Heatmap of the top 30 differentially expressed proteins between HCs (green) and thrPAPS (red), hierarchically clustered; rows scaled to row z-score. (D) Pathway enrichment (Metascape) of proteins upregulated in thrPAPS; bars coloured by –log₁₀(P). Complement and coagulation cascades, platelet degranulation and acute-phase response dominate the upregulated programme.

### A three-protein panel has strong discriminatory performance for thrPAPS

To identify coordinated protein expression patterns associated with thrPAPS, we performed WGCNA on the 1,447 plasma proteins retained after quality control. This analysis identified 11 co-expression modules, of which the brown (n=178 proteins) and green (n=113 proteins) modules showed the strongest positive associations with disease status (r=0.73 and r=0.60, respectively; **Supplementary Table 8**; **Figure 3A**).

**Figure 3.**
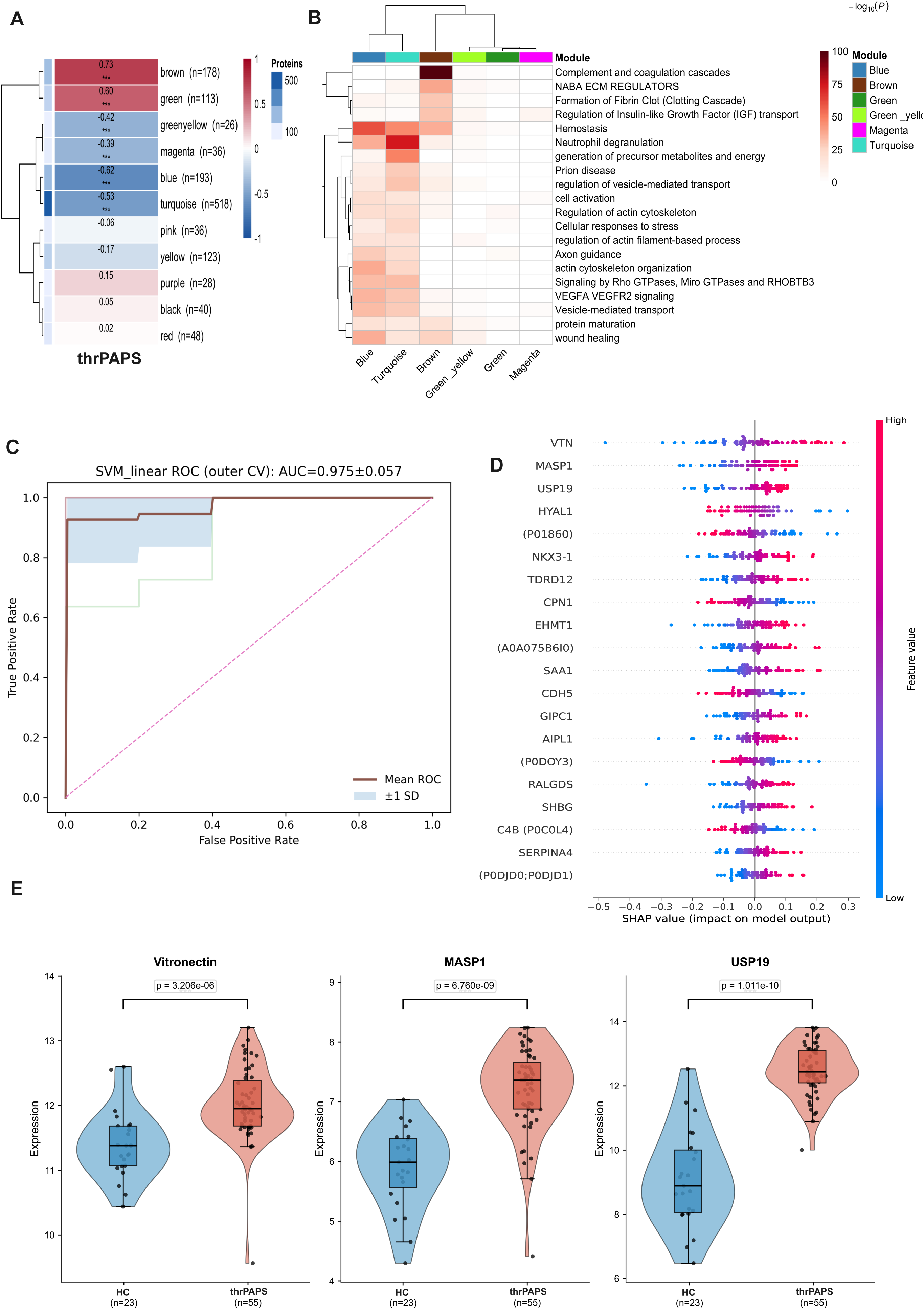
WGCNA-derived plasma proteins provide a minimal diagnostic classifier for thrPAPS. (A) Module–trait relationship heatmap from WGCNA of 1,447 plasma proteins, showing Pearson correlation between each module eigengene and thrPAPS status; numbers of proteins per module are in parentheses. The brown (n=178, r=0.73) and green (n=113, r=0.60) modules showed the strongest positive correlation with disease. (B) Pathway enrichment per module; the brown module is enriched for complement and coagulation cascades, NABA ECM regulators and fibrin-clot formation. (C) Receiver operating characteristic (ROC) curve for the linear SVM classifier trained on the 178 brown-module proteins under 5-fold nested cross-validation (mean AUC = 0.975 ± 0.057); shaded area = ±1 SD. (D) SHAP beeswarm plot of the top 20 features driving SVM predictions, ordered by mean absolute SHAP value. Vitronectin (P04004), MASP1 (P48740) and USP19 (O94966) are the highest-ranked predictors. (E) Violin/box plots of the three top-ranked proteins (Vitronectin, MASP1, USP19) in HCs (n=23) vs thrPAPS (n=55), all significantly elevated in thrPAPS (Wilcoxon P values indicated).

Pathway enrichment analysis showed that the brown module was enriched for complement and coagulation cascades, extracellular matrix regulators (NABA ECM regulators), and fibrin clot formation. In contrast, the green module showed no specific functional enrichment (**Figure 3B**). Given its strong disease association and biological relevance, we evaluated the diagnostic potential of the 178 proteins in the brown module using a machine-learning classification model. Using nested cross-validation (5-fold outer, 3-fold inner), we benchmarked elastic-net logistic regression, random forests, linear support vector machines (SVMs), and XGBoost. The linear SVM achieved the highest performance, with a mean AUROC of 0.975 ± 0.057 across outer folds, and was, therefore, selected for all subsequent analyses (**Supplementary Table 9, Figure 3C**).

To identify the proteins most contributing to disease classification, SHAP (SHapley Additive exPlanations) values (31) were computed for the final SVM model trained on the full dataset (**Figure 3D)**. The highest-ranking proteins were Vitronectin, followed by Mannan-binding lectin serine protease 1 (MASP-1) and Ubiquitin carboxyl-terminal hydrolase 19 (USP19). For most top-ranked proteins, high protein abundance (red) was associated with positive SHAP values, indicating a contribution towards classification as thrPAPS. For example, all three of the highest-ranking proteins showed statistically significant increase in abundance between disease and HC (Vitronectin p = 3.21 × 10 ⁶, MASP1 p = 6.76 × 10⁹ and USP19 p = 1.01 × 10^10^; **Figure 3E**).

To assess whether a reduced set of proteins could maintain classification performance, SHAP-ranked protein subsets of increasing size (top 3, 5, 10, 15, and 20) were evaluated using the same nested cross-validation framework with the linear SVM (**Supplementary Table 9**). A three-panel achieved a mean AUROC of 0.989, exceeding that of the full 178-protein panel (AUROC = 0.975). All reduced panels (top 3 through top 20) achieved mean AUROCs between 0.989 and 1.000, with narrow standard deviations, indicating that the discriminative information was concentrated in a small number of key proteins.

### Stratified analyses across high-risk thrPAPS phenotypes

Across arterial, venous, and mixed arterial/venous thrombosis subgroups, 61% of upregulated proteins were shared (**Figure 4A**), indicating a conserved thromboinflammatory signature independent of thrombosis location. However, the mixed arterial/venous phenotype showed the greatest degree of subgroup-specific dysregulation, including proteins involved in coagulation, fibrinolysis, and complement pathways (e.g. CFD, F9, CFHR4, FGA), consistent with more extensive molecular perturbation (**Supplementary Figure 2**).

**Figure 4.**
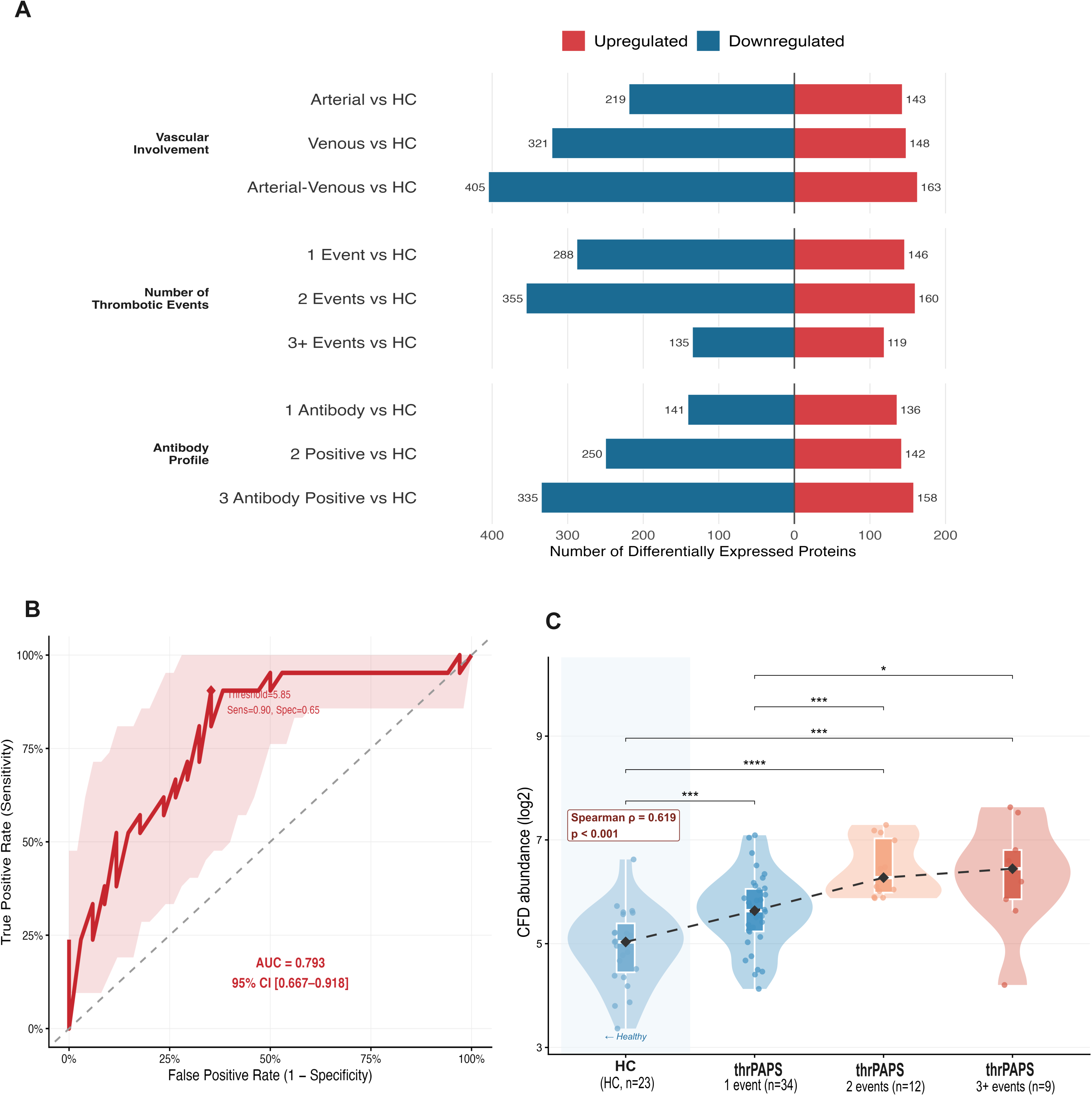
Plasma proteomic stratification of thrPAPS by severity and antibody burden. (A) Bar chart showing the numbers of upregulated (red) and downregulated (blue) proteins in each thrPAPS subgroup versus HCs: by vascular involvement (arterial, venous, arterial–venous), by number of thrombotic events (1, 2, ≥3), and by aPL profile (single-, double-, triple-positive). (B) ROC curve for plasma CFD discriminating single-event from recurrent APS (≥2 events); AUC = 0.793 (95% CI 0.667–0.918); optimal threshold 5.85, giving sensitivity 0.90 and specificity 0.65. (C) CFD abundance (log₂) across HC, APS with 1 thrombotic event (n=34), 2 events (n=12), and ≥3 events (n=9). Dashed line indicates Spearman ρ = 0.619, P < 0.001 between CFD and thrombotic event count; BH-adjusted pairwise P values shown.

Stratification by thrombotic event number revealed a progressive expansion of proteomic dysregulation (Figure 4A). Patients with ≥3 events showed distinct upregulation of complement- and coagulation-related proteins. In comparison, patients with 2 and ≥3 events shared a set of proteins including CFD, EFEMP1, MASP2, and C4BPB, consistent with a recurrence-associated molecular programme (Supplementary Table 10). In a targeted comparison of recurrent (≥2 events) versus single-event disease accounting for age, sex and treatments as covariates, complement factor D (CFD) was the top-ranked differentially abundant protein (LFC >0.5 and p-value <0.001). CFD abundance correlated with thrombotic event count (r=0.619, p<0.001) and discriminated recurrent from single-event disease (AUC=0.793; Figure 4B–C), supporting a role for alternative complement pathway activation in thrombotic burden and recurrence risk.

Stratification by aPL profile showed a progressive increase in the number of differentially expressed proteins with increasing aPL burden, from 276 in single aPL-positive patients to 493 in triple-positive patients (**Figure 4A**). A shared set of 100 upregulated proteins (54%) was observed across all groups, indicating a common aPL-associated signature. In contrast, double-and triple-aPL positive patients showed additional upregulation of coagulation-related proteins, including fibrinogen chains, and a 44-protein signature enriched for fibrinolysis, complement activation, and clot formation pathways (**Supplementary Figure 2-3, Supplementary Table 10**).

### A latent factor tracks aPL burden and maps to an innate immune-receptor signalling network

To resolve the principal axes of molecular variation in thrPAPS, we applied MOFA2 to jointly model plasma proteomics (980 proteins) and whole-blood transcriptomics (5,000 most variable genes). After model fitting and pruning of factors explaining less than 1% of variance in both views, 13 latent factors were retained, accounting for 55.8% of proteomic and 35.9% of transcriptomic variance overall (**Figure 5A**). Each factor captured a distinct proportion of variance across the proteomic and transcriptomic views, with some factors shared across both modalities and others largely view-specific (**Figure 5B**).

**Figure 5.**
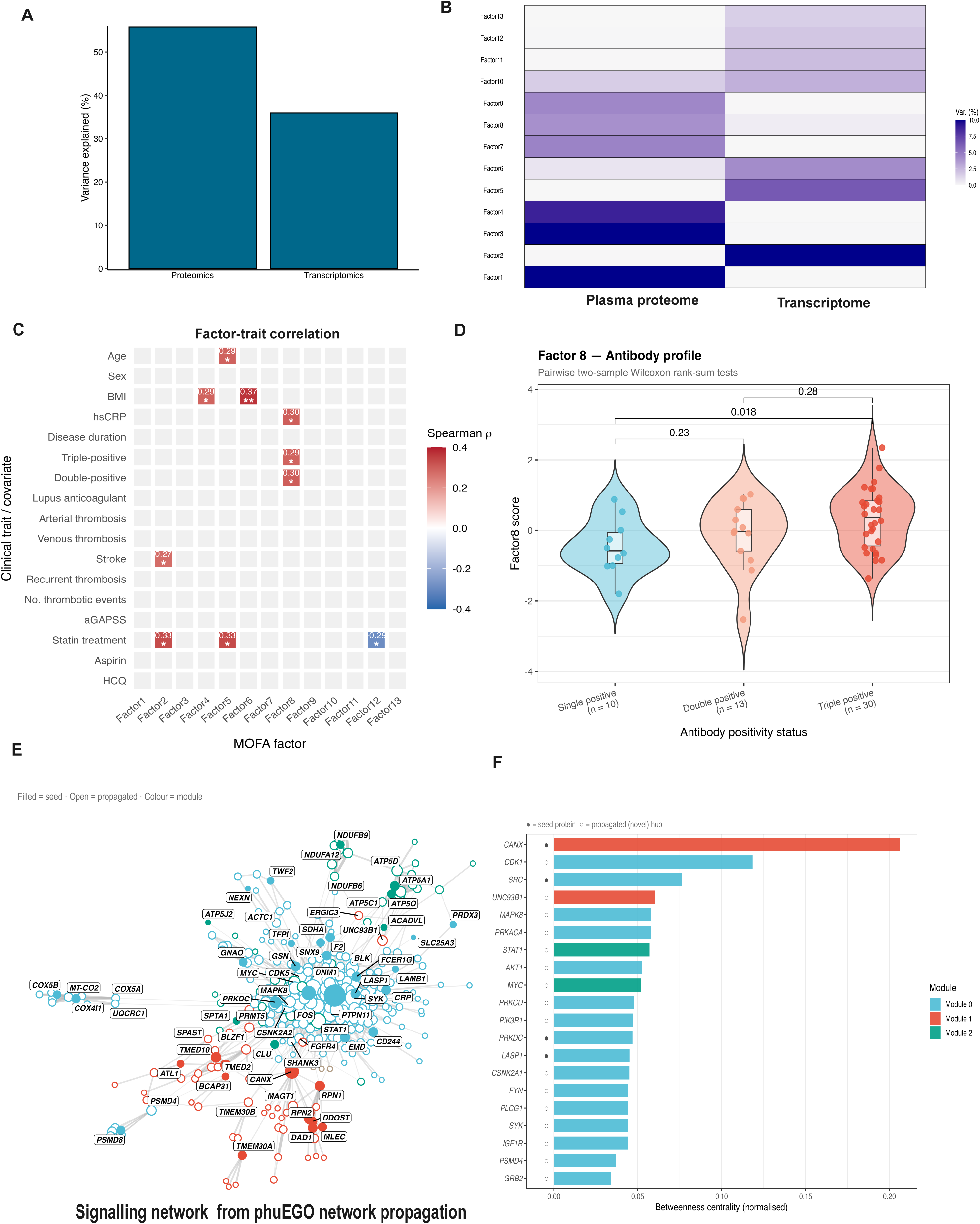
A complement-rich latent factor (Factor 8) is associated with antiphospholipid antibody burden and embeds within immune-receptor signalling networks. (A) Total variance explained (%) by the MOFA model in each input view (plasma proteome and whole-blood transcriptome), summed across all 13 factors. (B) Per-factor variance explained (%) decomposed by view. Each factor (rows, Factor1–Factor13) is shown for the plasma proteome and transcriptome (columns); colour intensity denotes the percentage of variance that factor explains within each view (note the colour scale is capped at 10% and is not directly comparable to the totals in panel A). (C) Factor–trait association screen. (D) Factor 8 score across antiphospholipid antibody positivity strata (single-positive, n = 10; double-positive, n = 13; triple-positive, n = 30). Violin plots show the score distribution; overlaid box plots denote the median and interquartile range. Brackets show pairwise two-sided Wilcoxon rank-sum p-values; Factor 8 score increases with antibody burden (single vs triple p = 0.018). (E) Signalling network derived from phuEGO network propagation seeded with the top 100 Factor 8 positive proteins. Filled nodes are input seeds; open nodes are recovered by propagation; node colour denotes the assigned network module (legend), with inter-module nodes shown separately (F). Bar plot showing the top 20 hub genes in the propagated network, ranked by betweenness centrality. recovered by propagation; node colour denotes the assigned network module (legend), with inter-module nodes shown separately (F). Bar plot showing the top 20 hub genes in the propagated network, ranked by betweenness centrality.

In the factor–covariate screen, Factor 8 was associated with double- and triple-aPL positivity, and hsCRP (**Figure 5C**). Factor 8 score was significantly higher in triple-positive than in non-triple-aPL positive patients (Wilcoxon p = 0.036) and increased monotonically across single-, double-, and triple-aPL positivity (Spearman rho = 0.33, p = 0.017), indicating a graded association with aPL burden (**Supplementary Figure 4, Figure 5 C-D)**. The proteins driving Factor 8 included terminal complement proteins (C6, C7, C8B, C8G, C9, CFB), platelet- and coagulation-associated proteins (F2, F11, TFPI, TBXAS1, PTGS1) and acute-phase proteins (CRP, SAA1, SAA4). Gene-set enrichment of the ranked proteomic loadings (fgsea; Reactome and Hallmark collections) identified significant enrichment of the complement cascade (NES = +2.11, padj = 0.049) and plasma lipoprotein remodelling (NES = +2.25, padj = 0.033) (**Supplementary Table 11**). Despite explaining less transcriptomic variance, factor 8 gene loadings were strongly enriched for type I interferon response (HALLMARK_INTERFERON_ALPHA_RESPONSE, NES = 2.64, padj = 3.6 × 10⁻⁹), type II interferon response (HALLMARK_INTERFERON_GAMMA_RESPONSE, NES = 2.58, padj = 3.6 × 10⁻⁹), and heme metabolism (NES = 2.32, padj = 6.7 × 10⁻⁸, **Supplementary Table 11)**. To place this complement-rich plasma signature in a signalling network context, we used the top 100 proteins contributing to Factor 8 as seed nodes for network propagation with phuEGO. This resolved three functional modules (**Figure 5E; Supplementary Table 12**). Pathway enrichment analysis showed that module 0 was enriched for immune cell signalling, module 1 for asparagine N-linked glycosylation, and module 2 for mitochondrial oxidative phosphorylation. Among the top ten hubs ranked by betweenness centrality were CANX, CDK1, SRC, UNC93B1, MAPK8, PRKACA, STAT1, AKT1, MYC, and PRKCD (**Figure 5F**). These findings place Factor 8 within a broader thrombo-inflammatory network spanning innate immune receptor signalling, glycosylation, and mitochondrial metabolism.

### Reactive oxygen species-associated proteomic signatures are key indicators of disease activity in thrPAPS

To examine plasma protein signatures associated with disease activity, we performed longitudinal profiling from 9 patients with available samples from inactive (>9 months after a thrombotic event) and active (at the time of a new thrombotic event) states (**Figure 6A**). Paired differential abundance analysis identified 76 proteins upregulated and 18 downregulated in the active state (|log₂FC|>1, p<0.05; **Figure 6B; Supplementary Table 13**).

**Figure 6.**
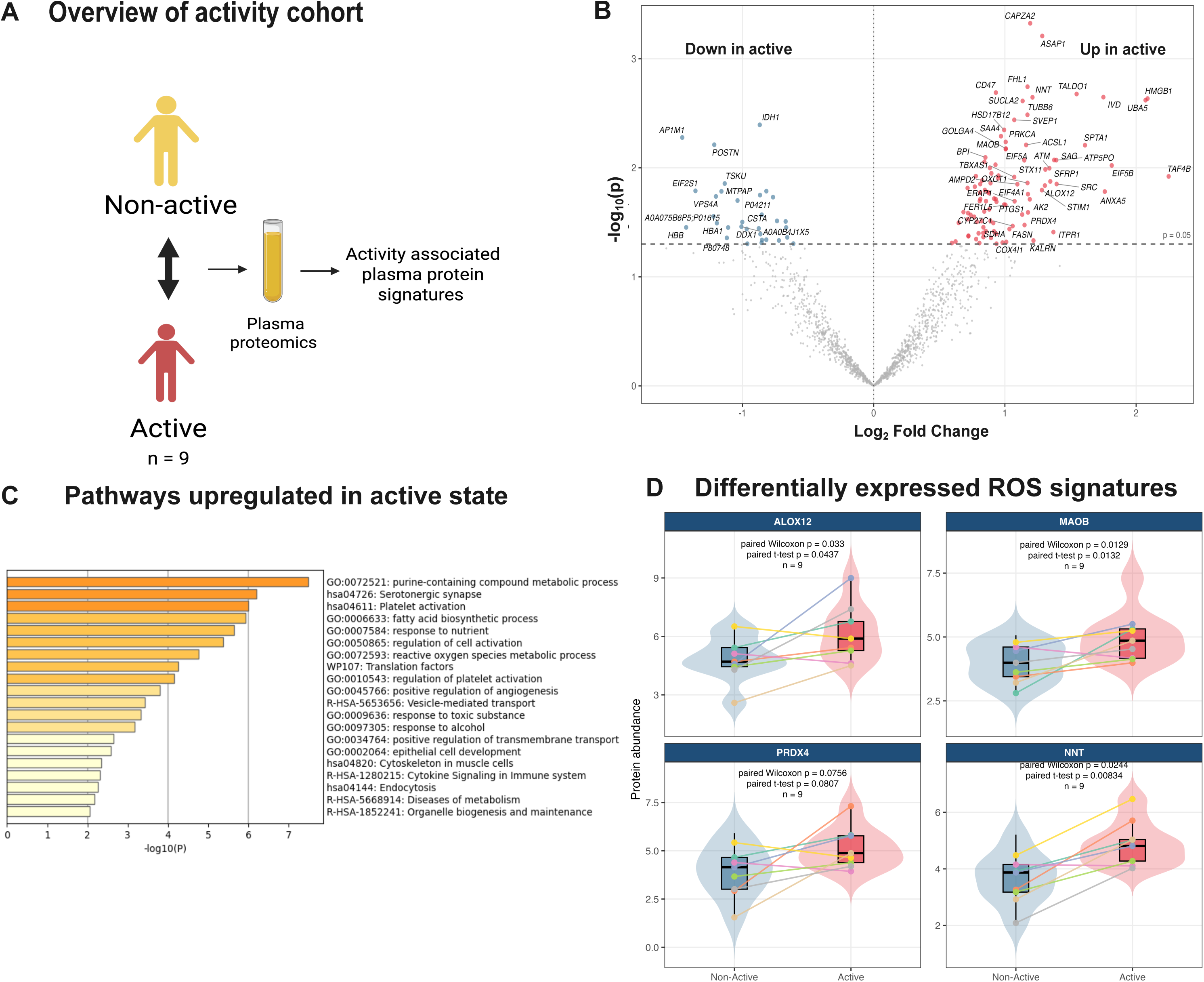
Reactive oxygen species–associated proteomic signatures distinguish active from non-active disease. **(A)** Schematic of the longitudinal activity cohort: nine patients with paired plasma proteomes collected during active and non-active states. **(B)** Volcano plot of paired limma differential expression (active vs non-active); 76 proteins upregulated and 18 downregulated in the active state (|log₂FC| > 1, P < 0.05). **(C)** Metascape pathway enrichment of proteins upregulated in active disease **(D)** Paired violin/box plots of four ROS-related proteins (ALOX12, MAOB, PRDX4, NNT) in matched non-active and active samples (n=9); paired Wilcoxon and paired t-test P values shown. Coloured lines connect the same patient across the two disease states.

Upregulated proteins were enriched for platelet activation, metabolic and oxidative pathways, including purine-containing compound metabolism and reactive oxygen species (ROS) metabolism (**Figure 6C**). Several upregulated proteins are directly involved in ROS production and redox regulation, including ALOX12, MAOB, PRDX4, and NNT (**Figure 6D**).

## Discussion

A major unmet need in thrPAPS is the molecular stratification of disease phenotypes linked to a high risk of poor outcomes, including recurrent thrombosis and multiple aPL positivity. Using integrated proteomic and transcriptomic profiling, we identified complement activation as the dominant molecular programme associated with thrombosis recurrence and increasing aPL burden. Longitudinal analyses further linked disease activity to oxidative stress pathways, while multi-omics integration connected circulating thromboinflammatory signatures to interferon-associated transcriptional programmes. Together, these findings provide a systems-level framework for understanding high-risk APS and identify molecular pathways with potential relevance for biomarker development and targeted therapeutic intervention.

The consistency of complement activation signatures across differential abundance, co-expression network, and latent factor analyses suggests that complement activation is a central component of the thromboinflammation cascade in APS. aPL-mediated complement activation has been described via both the classical and alternative pathways (32,33). Complement-related proteins were enriched in recurrent thrombosis phenotypes. Complement factor D (CFD) emerged as a marker of recurrence and thrombotic burden. As the rate-limiting enzyme of the alternative complement pathway, CFD drives the pathway amplification loop (34). Previous studies have linked terminal complement activation and C5b-9 deposition to severe APS manifestations, including catastrophic and recurrent thrombosis (35), while elevated Bb fragment levels support a role for alternative pathway activation in high-risk disease (36). We also observed enrichment of platelet and acute-phase response proteins, mainly serpins, which regulate coagulation, fibrinolysis, and innate immunity (37). These findings are consistent with a recent proteomic study implicating coagulation, platelet activation and innate immunity pathways in APS (19). Given the emergence of complement inhibitors in APS and related thromboinflammatory disorders, CFD may warrant further evaluation as both a biomarker of recurrence risk and a marker of alternative pathway activation.

Platelet activation is increasingly recognised as a key contributor to thromboinflammation in APS, and recent studies have shown that platelet–neutrophil aggregates promote NET formation, while inhibition of CD40–CD40L signalling can attenuate the platelet–neutrophil/NET axis (38). Consistent with these observations, a three-protein classifier comprising Vitronectin, MASP1, and USP19 (39) highlighted the convergence of platelet, complement, and autophagy-related pathways in distinguishing thrPAPS from healthy controls.

Transcriptomic analysis within the thrPAPS cohort identified an interferon-associated co-expression module that was positively associated with triple aPL positivity. These findings extend previous reports from our group and others demonstrating activation of both type I and type II interferon pathways in primary APS (12–17). While interferon signatures have consistently emerged from whole-blood transcriptomic studies, their relationship to circulating protein signatures has remained unclear. Multi-omics integration provided a complementary and unbiased perspective, identifying a latent factor whose score increased progressively across single-, double-, and triple-aPL positivity. This graded relationship is particularly relevant because aPL burden is itself among the strongest predictors of thrombosis and recurrence risk in APS (3–5). Factor 8 was characterised by complement, coagulation, and platelet-associated proteins, as well as acute-phase reactants, while enrichment analysis of the ranked factor loadings confirmed the over-representation of the complement cascade. The convergence of differential abundance, co-expression and latent factor analyses therefore supports complement activation as a defining feature of high-risk thrPAPS and suggests that complement and interferon pathways represent interconnected components of disease severity.

Network propagation placed this complement-rich factor within a broader signalling context and identified several hubs previously implicated in APS pathogenesis.. SRC and PI3K–AKT play a key role in platelet activation (40). In APS, Anti-β2GP1 antibodies enhance platelet activation and upregulate the mTORC2/AKT pathway (41). STAT1 and PRKCD provide links to interferon-driven immune activation consistent with the interferon-associated transcriptomic programme identified here; notably, PRKCD has established roles in B-cell tolerance and interferon signalling, positioning it at the intersection of thromboinflammatory pathways (13,42,43). Additional hubs, including MAPK8, PRKACA and MYC, further support the involvement of stress-response, immune-regulatory and transcriptional programmes within the propagated network. UNC93B1, a regulator of nucleic acid-sensing Toll-like receptors, is also consistent with innate immune activation associated with NET formation and interferon induction (44,45), whereas CANX highlights the importance of glycosylation and protein-trafficking pathways in the processing of complement proteins and immunoglobulins (46,47).

Mitochondrial oxidative phosphorylation dysfunction is increasingly recognised as a driver of inflammation and endothelial damage in autoimmune disorders. Studies have shown that aPL binding to monocyte and neutrophil membranes alters mitochondrial membrane potential and hyperactivates the electron transport chain (48), leading to oxidative stress and a proinflammatory, prothrombotic state in APS (49). Consistent with these observations, longitudinal sample profiling identified altered redox metabolism and ROS-associated proteins during active disease, supporting a role for oxidative stress in mediating disease activity in thrPAPS (50). Upregulated proteins in the active state were also enriched for platelet activation, with some of them also contributing to the platelet-neutrophil-NETs axis. ALOX12 (arachidonate 12-lipoxygenase) generates 12-HETE, a lipid mediator that primes neutrophils for NETosis and enhances platelet–neutrophil interactions (51). NNT (nicotinamide nucleotide transhydrogenase), a mitochondrial inner membrane enzyme essential for NADPH regeneration, is consistent with mitochondrial membrane disruption during NETosis (52).

Together, these findings connect circulating biomarkers to intracellular pathways through which aPL activate a thromboinflammation loop by engaging interconnected innate immune system components, including complement, the interferon-STAT-1 axis, platelets, neutrophils/ NETs, and identify potential targets for future treatments.

Strengths of the study include the integration of proteomics, transcriptomics, machine learning and longitudinal sampling in a relatively large sample, given the rarity of the disease, of well-characterised patients with exclusively primary APS and a thrombotic history. Limitations include a predominantly White population, small sample sizes in some subgroup analyses, and the absence of an independent validation cohort. However, paired plasma proteomic and whole-blood transcriptomic datasets in primary thrombotic APS are not currently available elsewhere, reflecting both the rarity of the disease and the novelty of this resource. The consistency of findings across multiple analytical approaches supports the robustness of the identified molecular programmes.

In conclusion, our study provides an integrated systems-level view of high-risk thrPAPS and identifies complement activation as a central molecular axis linking interferon and platelet/coagulation-associated pathways. These findings support the development of complement-informed and network-guided therapeutic strategies that extend beyond anticoagulation alone and may ultimately enable more precise risk stratification and treatment selection in APS.

## Author contributions

SS: Transcriptomic, Proteomic and Multi-Omic analysis, Methodology, Data Curation, Data interpretation, Visualization, Writing – original draft, Writing – review & editing.

MB and K-MV: Transcriptomic data generation and analysis, Data Curation, Writing – review & editing.

GR: Proteomic data generation, Data curation, Writing – review & editing. PS: Resources, Writing – review & editing.

MS: Proteomic data generation, Data curation, Resources, Writing – review & editing.

EP: Supervision of Bioinformatic analysis, Methodology, Resources, Writing – original draft, Writing – review & editing.

MGT: Conceptualization-Study design, Project Supervision, Methodology, Resources, Patient recruitment, clinical data and sample collection, Funding acquisition, Data interpretation, Writing – original draft, Writing – review & editing.

## Data availability statement

The complete RNA-sequencing dataset, including raw counts and normalised expression matrices, has been deposited in the Gene Expression Omnibus (GEO) database under accession number GSE205465. The mass spectrometry plasma proteomics data have been deposited in the ProteomeXchange Consortium via the PRIDE partner repository (Project accession: PXD080198). The code used for analysis is available upon reasonable request.

## Ethics statement

The studies involving humans were approved by Ethical Committee of the Laiko Hospital, Athens, Greece (number 509). The studies were conducted in accordance with the local legislation and institutional requirements. The participants provided their written informed consent to participate in this study.

## Acknowledgments

Transcriptomic analysis was supported by the project “The Greek Research Infrastructure for Personalised Medicine (pMedGR)” (MIS 5002802) which is implemented under the Action ‘Reinforcement of the Research and Innovation Infrastructure”, funded by the Operational Programme “Competitiveness, Entrepreneurship and Innovation” (NSRF 2014–2020) and co-financed by Greece and the European Union (European Regional Development Fund).

Proteomic analysis was supported by the National Recovery and Resilience Plan “Greece 2.0” - Next Generation EU Grant ID: 5173820 Duration:2023 - 2025 Type: RRF Action 16624 - Creation, Expansion and Upgrading of Research Centres (GSRI) infrastructure Title: Upgrading of the existing building and infrastructure of the Biomedical Sciences Research Center Alexander Fleming. This project has been made possible in part by grant number 2024-351060 from the Chan Zuckerberg Initiative DAF, an advised fund of Silicon Valley Community Foundation to SS and EP. SS, and EP are supported by EMBL and EMBL-EBI with core funds. EMBL IT is acknowledged for providing computer and data storage servers. Figure 1 was created with Biorender.com

## References

1. Barbhaiya M, Zuily S, Naden R, Hendry A, Manneville F, Amigo MC, et al. 2023 ACR/EULAR antiphospholipid syndrome classification criteria. Ann Rheum Dis. 2023 Oct 1;82(10):1258–70. doi:10.1136/ard-2023-224609

2. Tektonidou MG, Andreoli L, Limper M, Amoura Z, Cervera R, Costedoat-Chalumeau N, et al. EULAR recommendations for the management of antiphospholipid syndrome in adults. Ann Rheum Dis. 2019 Oct 1;78(10):1296–304. doi:10.1136/annrheumdis-2019-215213

3. Pengo V, Ruffatti A, Legnani C, Gresele P, Barcellona D, Erba N, et al. Clinical course of high-risk patients diagnosed with antiphospholipid syndrome. J Thromb Haemost. 2010 Feb 1;8(2):237–42. doi:10.1111/j.1538-7836.2009.03674.x

4. Ahmadzadeh Y, Magder LS, de Andrade DCO, Paredes-Ruiz D, Tektonidou MG, Pengo V, et al. Predictors of Mortality in Antiphospholipid Antibody–Positive Patients: Prospective Results From Antiphospholipid Syndrome Alliance for Clinical Trials and International Networking Clinical Database and Repository. Arthritis Care Res. 2025;77(6):760–6. doi:10.1002/acr.25503

5. Thaler J, Parides M, de Andrade DCO, Ruiz DP, Tektonidou MG, Pengo V, et al. Clinical and biologic predictors of thrombosis in persistently antiphospholipid antibody-positive patients: Prospective analysis of the International APS ACTION Clinical Database and Repository (‘Registry’). Ann Rheum Dis. 2026 Mar 1;85(3):497–506. doi:10.1016/j.ard.2025.10.019

6. Cervera R, Serrano R, Pons-Estel GJ, Ceberio-Hualde L, Shoenfeld Y, de Ramón E, et al. Morbidity and mortality in the antiphospholipid syndrome during a 10-year period: a multicentre prospective study of 1000 patients. Ann Rheum Dis. 2015 Jun;74(6):1011–8. doi:10.1136/annrheumdis-2013-204838

7. Raschi E, Borghi MO, Tedesco F, Meroni PL. Antiphospholipid syndrome pathogenesis in 2023: an update of new mechanisms or just a reconsideration of the old ones? Rheumatology. 2024 Feb 1;63(SI):SI4–13. doi:10.1093/rheumatology/kead603

8. Knight JS, Branch DW, Ortel TL. Antiphospholipid syndrome: advances in diagnosis, pathogenesis, and management. BMJ. 2023 Feb 27;380:e069717. doi: 10.1136/bmj-2021-069717.

9. Tektonidou MG. Cardiovascular disease risk in antiphospholipid syndrome: Thrombo-inflammation and atherothrombosis. J Autoimmun. 2022 Apr 1;128:102813. doi:10.1016/j.jaut.2022.102813

10. McDonnell T, Wincup C, Buchholz I, Pericleous C, Giles I, Ripoll V, et al. The role of beta-2-glycoprotein I in health and disease associating structure with function: More than just APS. Blood Rev. 2020 Jan 1;39:100610. doi:10.1016/j.blre.2019.100610

11. Lopez-Pedrera C, Pérez-Sánchez C, Tektonidou MG. Towards precision medicine in antiphospholipid syndrome. Lancet Rheumatol. 2025 Aug 1;7(8):e576–89. doi:10.1016/S2665-9913(25)00094-3

12. Verrou KM, Sfikakis PP, Tektonidou MG. Whole blood transcriptome identifies interferon-regulated genes as key drivers in thrombotic primary antiphospholipid syndrome. J Autoimmun. 2023 Jan 1;134:102978. doi:10.1016/j.jaut.2022.102978

13. Baltsiotis M, Verrou KM, Sfikakis PP, Tektonidou MG. RNA sequencing-derived gene co-expression and drug-gene interaction analysis reveal STAT1 as a potential therapeutic target in thrombotic antiphospholipid syndrome. Front Immunol. 2026 Mar 5;17. doi:10.3389/fimmu.2026.1741872

14. van den Hoogen LL, Fritsch-Stork RDE, Versnel MA, Derksen RHW, van Roon JAG, Radstake TRD. Monocyte type I interferon signature in antiphospholipid syndrome is related to proinflammatory monocyte subsets, hydroxychloroquine and statin use. Ann Rheum Dis. 2016 Dec 1;75(12):e81. doi:10.1136/annrheumdis-2016-210485

15. Palli E, Kravvariti E, Tektonidou MG. Type I Interferon Signature in Primary Antiphospholipid Syndrome: Clinical and Laboratory Associations. Front Immunol. 2019 Mar 15;10. doi:10.3389/fimmu.2019.00487

16. Xourgia E, Tektonidou MG. Type I interferon gene expression in antiphospholipid syndrome: Pathogenetic, clinical and therapeutic implications. J Autoimmun. 2019 Nov;104:102311. doi: 10.1016/j.jaut.2019.102311.

17. Tektonidou MG, Verrou KM, Gakiopoulou H, Manoloukos M, Lembessis P, Hatzis P, et al. Kidney whole-transcriptome profiling in primary antiphospholipid syndrome reveals complement, interferons and NETs-related gene expression. Rheumatology. 2024 Nov 1;63(11):3184–90. doi:10.1093/rheumatology/keae397

18. Ambati A, Ma F, Kmetova K, Navaz S, Hoy CK, Sarosh C, et al. Molecular Stratification of Antiphospholipid Syndrome Through Integrative Analysis of the Whole-Blood RNA Transcriptome. Arthritis Rheumatol. 2026;78(5):1124–33. doi:10.1002/art.70021

19. Pine A, Butt A, Andreoli L, Knight JS, Gerosa M, Cecchi I, et al. A proteomic map of thromboinflammatory signatures in antiphospholipid syndrome: results from antiphospholipid syndrome alliance for clinical trials and international networking (APS ACTION) registry. Front Immunol. 2025 Oct 16;16. doi:10.3389/fimmu.2025.1676578

20. Patro R, Duggal G, Love MI, Irizarry RA, Kingsford C. Salmon provides fast and bias-aware quantification of transcript expression. Nat Methods. 2017 Apr;14(4):417–9. doi:10.1038/nmeth.4197

21. Love MI, Huber W, Anders S. Moderated estimation of fold change and dispersion for RNA-seq data with DESeq2. Genome Biol. 2014 Dec 5;15(12):550. doi:10.1186/s13059-014-0550-8

22. McGurk KA, Dagliati A, Chiasserini D, Lee D, Plant D, Baricevic-Jones I, et al. The use of missing values in proteomic data-independent acquisition mass spectrometry to enable disease activity discrimination. Bioinformatics. 2020 Apr 1;36(7):2217–23. doi:10.1093/bioinformatics/btz898

23. Webel H, Niu L, Nielsen AB, Locard-Paulet M, Mann M, Jensen LJ, et al. Imputation of label-free quantitative mass spectrometry-based proteomics data using self-supervised deep learning. Nat Commun. 2024 Jun 26;15(1):5405. doi:10.1038/s41467-024-48711-5

24. Nielsen AB, Fjordside L, Drici L, Ottenheijm ME, Rasmussen C, Henningsson AJ, et al. The diagnostic potential of proteomics and machine learning in Lyme neuroborreliosis. Nat Commun. 2025 Oct 27;16(1):9322. doi:10.1038/s41467-025-64903-z

25. Ritchie ME, Phipson B, Wu D, Hu Y, Law CW, Shi W, et al. limma powers differential expression analyses for RNA-sequencing and microarray studies. Nucleic Acids Res. 2015 Apr 20;43(7):e47. doi:10.1093/nar/gkv007

26. Zhou Y, Zhou B, Pache L, Chang M, Khodabakhshi AH, Tanaseichuk O, et al. Metascape provides a biologist-oriented resource for the analysis of systems-level datasets. Nat Commun. 2019 Apr 3;10(1):1523. doi:10.1038/s41467-019-09234-6

27. Szklarczyk D, Gable AL, Lyon D, Junge A, Wyder S, Huerta-Cepas J, et al. STRING v11: protein–protein association networks with increased coverage, supporting functional discovery in genome-wide experimental datasets. Nucleic Acids Res. 2019 Jan 8;47(D1):D607–13. doi:10.1093/nar/gky1131

28. Langfelder P, Horvath S. WGCNA: an R package for weighted correlation network analysis. BMC Bioinformatics. 2008 Dec 29;9(1):559. doi:10.1186/1471-2105-9-559

29. Argelaguet R, Arnol D, Bredikhin D, Deloro Y, Velten B, Marioni JC, et al. MOFA+: a statistical framework for comprehensive integration of multi-modal single-cell data. Genome Biol. 2020 May 11;21:111. doi:10.1186/s13059-020-02015-1

30. Giudice G, Chen H, Koutsandreas T, Petsalaki E. phuEGO: A Network-Based Method to Reconstruct Active Signaling Pathways From Phosphoproteomics Datasets. Mol Cell Proteomics MCP. 2024 Apr 19;23(6):100771. doi:10.1016/j.mcpro.2024.100771

31. Lundberg S, Lee SI. A Unified Approach to Interpreting Model Predictions [Internet]. arXiv; 2017 [cited 2026 Jun 16]. Available from: http://arxiv.org/abs/1705.07874 doi:10.48550/arXiv.1705.07874

32. Oku K, Atsumi T, Bohgaki M, Amengual O, Kataoka H, Horita T, et al. Complement activation in patients with primary antiphospholipid syndrome. Ann Rheum Dis. 2009 Jun 1;68(6):1030–5. doi:10.1136/ard.2008.090670

33. Chaturvedi S, Braunstein EM, Brodsky RA. Antiphospholipid syndrome: Complement activation, complement gene mutations, and therapeutic implications. J Thromb Haemost. 2021 Mar 1;19(3):607–16. doi:10.1111/jth.15082

34. Barratt J, Weitz I. Complement Factor D as a Strategic Target for Regulating the Alternative Complement Pathway. Front Immunol. 2021 Sep 9;12. doi:10.3389/fimmu.2021.712572

35. Chaturvedi S, Braunstein EM, Yuan X, Yu J, Alexander A, Chen H, et al. Complement activity and complement regulatory gene mutations are associated with thrombosis in APS and CAPS. Blood. 2020 Jan 23;135(4):239–51. doi:10.1182/blood.2019003863

36. Yelnik CM, Lambert M, Drumez E, Martin C, Grolaux G, Launay D, et al. Relevance of inflammatory and complement activation biomarkers profiling in antiphospholipid syndrome patients outside acute thrombosis. Clin Exp Rheumatol. 2023 Sep;41(9):1875–1881. doi: 10.55563/clinexprheumatol/ric86c

37. Yaron JR, Zhang L, Guo Q, Haydel SE, Lucas AR. Fibrinolytic Serine Proteases, Therapeutic Serpins and Inflammation: Fire Dancers and Firestorms. Front Cardiovasc Med. 2021 Mar 25;8. doi:10.3389/fcvm.2021.648947

38. Naoum S, Spyropoulos C, Angelopoulou A, Gakiopoulou H, Katsimpoulas M, Gorgoulis VG, et al. CD40-CD40L inhibition attenuates platelet-neutrophil interaction and neutrophil extracellular trap release in primary antiphospholipid syndrome. Ann Rheum Dis. 2026 May 1;85(5):858–72. doi:10.1016/j.ard.2025.12.012

39. Jin S, Tian S, Chen Y, Zhang C, Xie W, Xia X, et al. USP19 modulates autophagy and antiviral immune responses by deubiquitinating Beclin-1. EMBO J. 2016 Apr 1;35(8):866–80. doi:10.15252/embj.201593596

40. Senis YA, Mazharian A, Mori J. Src family kinases: at the forefront of platelet activation. Blood. 2014 Sep 25;124(13):2013–24. doi:10.1182/blood-2014-01-453134

41. Tang Z, Shi H, Chen C, Teng J, Dai J, Ouyang X, et al. Activation of Platelet mTORC2/Akt Pathway by Anti-β2GP1 Antibody Promotes Thrombosis in Antiphospholipid Syndrome. Arterioscler Thromb Vasc Biol. 2023 Oct;43(10):1818–1832. doi: 10.1161/ATVBAHA.123.318978

42. Belot A, Kasher PR, Trotter EW, Foray AP, Debaud AL, Rice GI, et al. Protein Kinase Cδ Deficiency Causes Mendelian Systemic Lupus Erythematosus With B Cell-Defective Apoptosis and Hyperproliferation. Arthritis Rheum. 2013;65(8):2161–71. doi:10.1002/art.38008

43. Varinou L, Ramsauer K, Karaghiosoff M, Kolbe T, Pfeffer K, Müller M, et al. Phosphorylation of the Stat1 Transactivation Domain Is Required for Full-Fledged IFN-γ-Dependent Innate Immunity. Immunity. 2003 Dec 1;19(6):793–802. doi:10.1016/S1074-7613(03)00322-4

44. Pelka K, Bertheloot D, Reimer E, Phulphagar K, Schmidt SV, Christ A, et al. The Chaperone UNC93B1 Regulates Toll-like Receptor Stability Independently of Endosomal TLR Transport. Immunity. 2018 May 15;48(5):911–922.e7. doi:10.1016/j.immuni.2018.04.011

45. Sun X, Gui Y, Lu Y, Wang K, Yao J, Li Z, et al. Exploring potential biomarkers of NETosis-Related genes in spinal cord injury through machine learning and multi-omics analysis. Biochem Biophys Rep. 2026 Jan 20;45:102439. doi:10.1016/j.bbrep.2025.102439

46. Tsukamoto H, Tousson A, Circolo A, Marchase RB, Volanakis JE. Calnexin is associated with and induced by overexpressed human complement protein C2. Anat Rec. 2002;267(1):7–16. doi:10.1002/ar.10070

47. Liu T, Han J, Zhang R, Tang Z, Yi G, Gong W, et al. Characteristics of purified anti-β2GPI IgG N-glycosylation associate with thrombotic, obstetric and catastrophic antiphospholipid syndrome. Rheumatology. 2022 Mar 1;61(3):1243–54. doi:10.1093/rheumatology/keab416

48. Perez-Sanchez C, Ruiz-Limon P, Aguirre MA, Bertolaccini ML, Khamashta MA, Rodriguez-Ariza A, et al. Mitochondrial dysfunction in antiphospholipid syndrome: implications in the pathogenesis of the disease and effects of coenzyme Q10 treatment. Blood. 2012 Jun 14;119(24):5859–70. doi:10.1182/blood-2011-12-400986

49. López-Pedrera C, Barbarroja N, Jimenez-Gomez Y, Collantes-Estevez E, Aguirre MA, Cuadrado MJ. Oxidative stress in the pathogenesis of atherothrombosis associated with anti-phospholipid syndrome and systemic lupus erythematosus: new therapeutic approaches. Rheumatology. 2016 Dec 1;55(12):2096–108. doi:10.1093/rheumatology/kew054

50. Pappa M, Ntouros PA, Papanikolaou C, Sfikakis PP, Souliotis VL, Tektonidou MG. Augmented oxidative stress, accumulation of DNA damage and impaired DNA repair mechanisms in thrombotic primary antiphospholipid syndrome. Clin Immunol. 2023 Sep 1;254:109693. doi:10.1016/j.clim.2023.109693

51. Li C, Gao P, Zhuang F, Wang T, Wang Z, Wu G, et al. Inhibition of ALOX12–12-HETE Alleviates Lung Ischemia–Reperfusion Injury by Reducing Endothelial Ferroptosis-Mediated Neutrophil Extracellular Trap Formation. Research. 7:0473. doi:10.34133/research.0473

52. Regan T, Conway R, Bharath LP. Regulation of immune cell function by nicotinamide nucleotide transhydrogenase. Am J Physiol-Cell Physiol. 2022 Apr;322(4):C666–73. doi:10.1152/ajpcell.00607.2020

