## Supplementary Document for "Multi-omics and network propagation reveal latent innate immune programmes stratifying high-risk thrombotic primary antiphospholipid syndrome"

This file contains Supplementary Methods, Supplementary Table 1, an index of the supplementary data tables (Supplementary Tables 2–13, provided as separate spreadsheet files), and the Supplementary Figures 1-3

### **Supplementary Methods**

#### **Preprocessing of whole-blood RNA-sequencing data**

Raw FASTQ files were downloaded and assessed for quality using FastQC v0.12.1, and the results were aggregated with MultiQC v1.19. Adapters and low-quality bases were removed using fastp v0.22.0. Transcript-level quantification was performed with Salmon v1.10.3, which uses quasi-mapping with sequence-specific and GC bias correction. Reads were quantified against the human GRCh38 reference transcriptome using the comprehensive Ensembl cDNA reference (accessed 16 February 2026). Salmon was run in single-end mode with automatic library-type detection and bias correction enabled. Transcript-level abundance estimates were summarised to the gene level for differential expression analysis with DESeq2, and a variance-stabilising transformation (VST) was applied for clustering, enrichment, and multi-omics integration (Supplementary Table 2). Canonical haemoglobin transcripts were excluded prior to normalisation, in line with established practice in whole-blood transcriptomics, to limit library-composition bias driven by highly abundant erythroid transcripts.

#### **WGCNA of whole-blood transcriptomic data**

Weighted gene co-expression network analysis (WGCNA) was performed on VST-transformed RNA-seq data from patients with thrPAPS using the WGCNA R package. To reduce noise and improve network robustness, genes were filtered by variance, retaining the top 50% most variable genes across samples. After quality control to remove outlier samples and low-quality genes, a signed co-expression network was constructed using a soft-thresholding power selected to approximate scale-free topology. Co-expression modules were identified by hierarchical clustering and dynamic tree cutting (minimum module size 30 genes; merge cut height 0.25). Module eigengenes were correlated (Pearson) with high-risk clinical traits, including recurrent thrombosis, venous and arterial thrombosis, stroke, microvascular involvement, and antiphospholipid antibody (aPL) positivity status (single-, double-, and triple-positive).

#### **Sample preparation for LC-MS/MS analysis**

ENplus-BEADS (25 µL) were rinsed three times with 200 µL of ENplus-WASH buffer in the supplied reaction tubes; each wash consisted of mixing at 1300 rpm for 1 min, magnetic separation, and discarding of the supernatant. With beads held on the magnetic rack, 50 µL of ENplus-BIND buffer was added to each bead pellet, followed by 50 µL of plasma. Tubes were removed from the rack, mixed at 1300 rpm for 30 min at 30 °C, and the supernatant removed after magnetic separation. Bead–plasma complexes were washed three times with 100 µL of ENplus-BIND buffer (room temperature, 1300 rpm, 1 min per wash). Proteins were lysed by adding 40 µL of LYSE-BCT and incubating with agitation at 1300 rpm for 10 min at 60 °C. Enzymatic digestion was performed by first adding 300 µL of RESUSPEND to 10 µL of DIGEST-L and shaking (room temperature, 500 rpm, 10 min), then adding this activated mixture and incubating at 1300 rpm for 60 min at 37 °C. The reaction was quenched with 60 µL of STOP solution (1 min, room temperature). Samples were resuspended (room temperature, 1300 rpm, 1 min) and transferred to cartridges in waste tubes. Peptides were cleaned up by centrifugation, washing sequentially with 100 µL of WASH1 (room temperature, 1000 rcf, 1 min) and 100 µL of WASH2 (room temperature, 2250 rcf, 1 min), followed by two elutions with 75 µL of ELUTE buffer (room temperature, 1000 rcf, 1 min per elution) into a common collection tube. Eluates were vacuum-dried (SpeedVac), resuspended in 15 µL of LC-MS/MS loading buffer (0.2% n-dodecyl-β-D-maltoside, 2% acetonitrile, 0.1% formic acid) and sonicated. Peptide concentrations were quantified by NanoDrop and diluted to 10 ng/µL prior to injection.

#### **Machine-learning model performance**

Model performance was estimated using 5-fold nested cross-validation (5 outer × 3 inner folds, both stratified; random_state = 42). The inner loop performed hyperparameter optimisation via RandomizedSearchCV (30 iterations, scored by AUROC) and the outer loop provided unbiased estimates of generalisation performance. Reported metrics include mean AUROC, accuracy, sensitivity, specificity, positive predictive value, negative predictive value, and Matthew’s correlation coefficient (Supplementary Table 9). The best-performing model (linear SVM) was refitted on the full dataset using RandomizedSearchCV with 50 iterations. SHAP (SHapley Additive exPlanations) values were computed using the shap library (v0.50.0): a LinearExplainer was applied to the elastic-net logistic regression and linear SVM, and a TreeExplainer to the random forest and XGBoost models. A minimal protein panel was constructed by iteratively evaluating classifier performance on increasing SHAP-ranked protein subsets (top 3, 5, 10, 15, and 20 proteins) under the same nested cross-validation framework.

#### **MOFA2 training**

Multi-Omics Factor Analysis was performed with MOFA2 (v1.12) to jointly model two data views, plasma proteomics and whole-blood transcriptomics, across the 55 patients with paired measurements. Proteomic intensities were log-transformed, and whole-blood RNA-seq counts were variance-stabilised (DESeq2 VST). Sample identifiers were harmonised across the proteomic and transcriptomic coding systems before matching, and only patients with both omics layers were retained.

Before model fitting, both views were filtered to remove features expected to contribute predominantly to technical variance. From the transcriptomic view, we removed haemoglobin genes, cytoplasmic and mitochondrial ribosomal-protein genes (RPL/RPS/MRPL/MRPS) and mitochondrially encoded genes (MT-); from the proteomic view, we removed database-flagged contaminants, keratins, and haemoglobin subunits, with haemoglobin removal applied consistently to both views. After filtering, the transcriptomic view was reduced to the 5,000 most variable genes and the proteomic view to proteins above the 30th percentile of variance (980 proteins). Both views were scaled to unit variance (scale_views = TRUE).

MOFA2 was trained with a Gaussian likelihood for both views, an initial factorisation of 13 factors, slow convergence mode, and a fixed random seed (45) for reproducibility. Following training, factors explaining less than 1% of variance in both views were pruned. The final model retained 13 factors, accounting for 55.8% and 35.9% of variance in the proteomic and transcriptomic views, respectively. Variance explained per factor and per view was obtained from the trained model.

For each retained factor, genes and proteins were ranked by their MOFA2 feature loadings. Gene-set enrichment analysis was performed on the full ranked loading vectors using the fgsea package (v1.28.0), with Reactome and Hallmark gene sets from the Molecular Signatures Database (MSigDB) as references. Normalised enrichment scores were computed with correction for gene-set size, and significance was assessed using permutation-based p-values corrected for multiple testing with the Benjamini–Hochberg method (Supplementary Table 11).

#### **Factor–covariate association testing**

Associations between MOFA factors and clinical traits or covariates were assessed using Spearman’s rank correlation rather than the Pearson-based default to provide robustness against outliers and ordered categorical variables (e.g., aPL positivity strata). For ordered antibody-positivity groups, monotonic trends were additionally evaluated using the Jonckheere–Terpstra trend test. A nominal p ≤ 0.05 was used to flag associations in the exploratory factor–trait screen; all 13 factors were displayed to indicate the full set of comparisons.

#### **Network propagation and module analysis**

The top 100 positively weighted proteins for the factor of interest were used as seed nodes for network propagation with phuEGO on a human protein–protein interaction network. All downstream network analysis and visualisation were performed in R. Networks were represented as undirected weighted graphs using the igraph R package (https://r.igraph.org/), with nodes representing proteins and edge weights reflecting phuEGO confidence scores. The community structure identified by phuEGO using the Leiden algorithm was retained, and edges linking nodes in different modules were classified as inter-module edges. For each module, hub genes were ranked by betweenness centrality, and the top hubs are reported in Figure 5F. Module-level functional enrichment was assessed using overrepresentation analysis against Hallmark, Reactome, and Gene Ontology Biological Process gene sets.

### **Supplementary Tables**

**Supplementary Table 1.** Demographic and clinical characteristics of 55 patients with thrombotic primary antiphospholipid syndrome (thrPAPS) and 23 age- and sex-matched healthy controls. Values are n (%) unless otherwise indicated; aPL, antiphospholipid antibody; S.D., standard deviation.

| Patient characteristics | Thrombotic primary APS (N=55) | Healthy controls (N=23) |
| --- | --- | --- |
| Demographics | | |
| Female, n (%) | 36 (65.5%) | 16 (69.6%) |
| Mean age, years; mean (S.D.) (range) | 43.3 (12.63) (18, 69) | 43.3 (9.06) (27, 54) |
| Disease duration, years; mean (S.D.) (range) | 8.6 (7.79) (0, 28) | — |
| Thrombosis | | |
| Venous thrombosis only, n (%) | 25/55 (45.5%) | — |
| Arterial thrombosis only, n (%) | 17/55 (30.9%) | — |
| Venous and arterial thrombosis, n (%) | 11/55 (20.0%) | — |
| Recurrent thrombosis, n (%) | 23/55 (41.8%) | — |
| Antiphospholipid antibodies | | |
| Anticardiolipin antibodies (IgG or IgM) | 46/55 (83.6%) | — |
| Anticardiolipin antibodies, IgG | 29/55 (52.7%) | — |
| Anticardiolipin antibodies, IgM | 35/55 (63.6%) | — |
| Anti-β2-glycoprotein I (IgG or IgM) | 36/55 (65.5%) | — |
| Anti-β2-glycoprotein I, IgG | 24/55 (43.6%) | — |
| Anti-β2-glycoprotein I, IgM | 23/55 (41.8%) | — |
| Lupus anticoagulant | 44/53 (83.0%) | — |
| Single aPL positivity | 10/53 (18.9%) | — |
| Double aPL positivity | 43/53 (81.1%) | — |
| Double aPL positivity (excluding triple-positive) | 13/53 (24.5%) | — |
| Triple aPL positivity | 30/53 (56.6%) | — |
| Complement | | |
| C3 levels, mean (S.D.) (range) | 99.7 (22.94) (62, 162) | — |
| C4 levels, mean (S.D.) (range) | 18.2 (7.48) (2, 33) | — |
| Cardiovascular risk factors | | |
| Hypertension | 16/55 (29.1%) | — |
| Dyslipidaemia | 8/55 (14.5%) | — |
| Smoking, ever | 32/55 (58.2%) | — |
| Smoking, current | 19/55 (34.5%) | — |
| Smoking, pack-years; mean (S.D.) (range) | 11.2 (15.22) (0, 50) | — |
| Family history of coronary disease | 5/55 (9.1%) | — |
| Treatment | | |
| Vitamin K antagonists | 55/55 (100%) | — |
| Hydroxychloroquine | 20/55 (36.4%) | — |
| Aspirin | 28/55 (50.9%) | — |
| Statin | 9/55 (16.4%) | — |
| Antihypertensive drugs | 15/55 (27.3%) | — |

### **Supplementary Tables 2–13**

**Supplementary Tables 2–13 are provided as separate spreadsheet files accompanying the manuscript.** The content of each table is summarised below.

| Table | Description |
| --- | --- |
| Table 2 | Gene-level count matrix and variance-stabilising transformed (VST) whole-blood expression values used for downstream transcriptomic analyses. |
| Table 3 | log2 transformed intensities values of plasma proteins |
| Table 4 | Imputed plasma proteomic intensity matrix (1,447 proteins × 78 samples) generated with the PIMMS self-supervised deep-learning framework. |
| Table 5 | Differentially expressed genes in whole blood, thrPAPS versus healthy controls (DESeq2; \|log2FC\| > 1, adjusted p < 0.05). |
| Table 6 | Whole-blood transcriptomic WGCNA module–trait associations across high-risk phenotypes, including the interferon-enriched pink module. |
| Table 7 | Differentially abundant plasma proteins, thrPAPS versus healthy controls (limma, age- and sex-adjusted; \|log2FC\| > 1, adjusted p < 0.05). |
| Table 8 | Plasma proteomic WGCNA module–trait associations with disease status, including the disease-associated brown and green modules. |
| Table 9 | Machine-learning classifier performance under nested cross-validation, including full and SHAP-reduced (top 3–20) protein panels. |
| Table 10 | Stratified plasma differential abundance results by thrombosis location, thrombotic event count, and aPL burden. |
| Table 11 | MOFA Factor 8 gene-set enrichment (fgsea; Reactome and Hallmark) for the proteomic and transcriptomic loading vectors. |
| Table 12 | phuEGO propagated network for Factor 8: node–module assignments |
| Table 13 | Paired differential abundance results, active versus non-active disease (n = 9 patients; limma; \|log2FC\| > 1, p < 0.05). |

**Supplementary Figures**

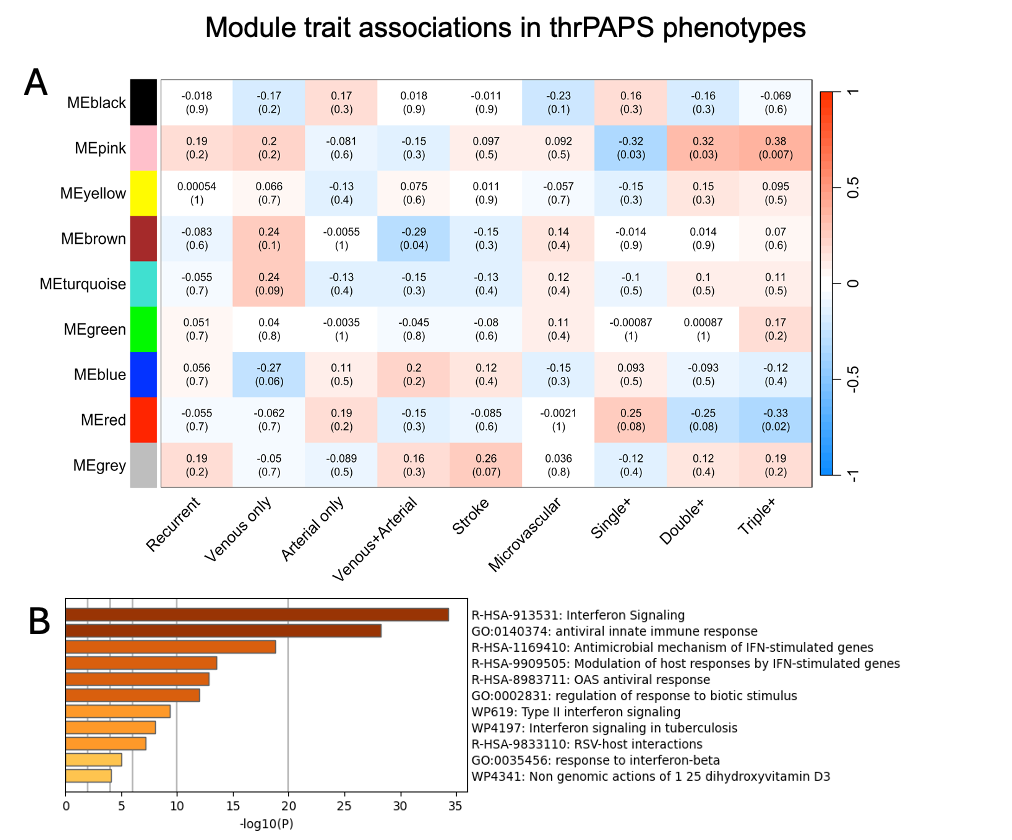

**Supplementary Figure 1.** WGCNA identifies an interferon-associated transcriptional module linked to high-risk thrPAPS phenotypes. (A) Weighted gene co-expression network analysis (WGCNA) of whole-blood transcriptomic data from patients with thrPAPS, showing module–trait associations across thrombotic manifestations and antiphospholipid antibody (aPL) burden phenotypes. The pink module is positively associated with triple-aPL positivity (r = 0.38, p = 0.007) and inversely associated with single-aPL positivity (r = −0.32, p = 0.03). (B) Metascape functional enrichment of genes within the pink module, demonstrating strong enrichment for interferon signalling, antiviral innate immune responses, interferon-stimulated gene pathways, and OAS-mediated antiviral responses, consistent with a type I interferon-driven transcriptional programme in high-risk thrPAPS.

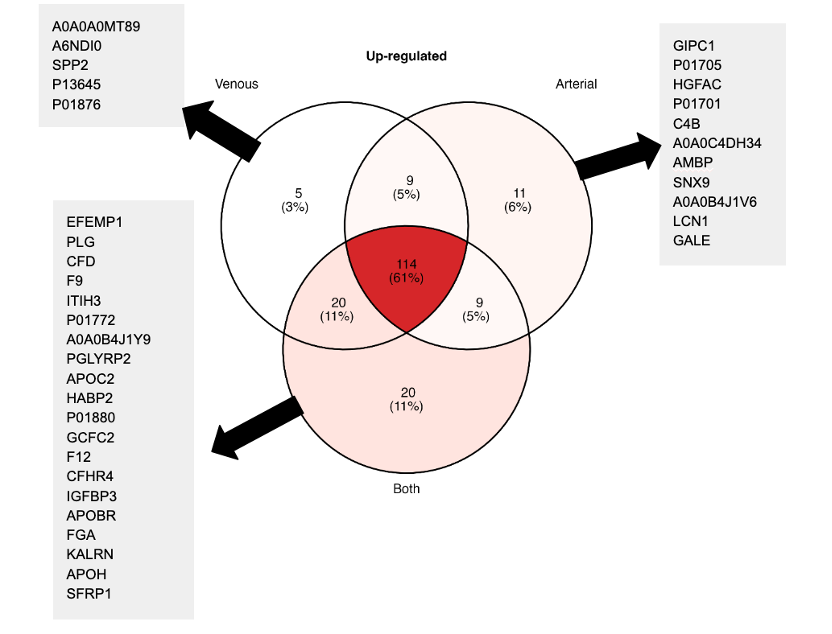

**Supplementary Figure 2.** Shared and phenotype-specific upregulated plasma proteins across thrombotic manifestations in thrPAPS. Venn diagram showing common and unique upregulated plasma proteins in venous, arterial, and combined venous + arterial thrombotic phenotypes compared with healthy controls. The mixed arterial/venous phenotype shows the greatest degree of subgroup-specific dysregulation, including coagulation-, fibrinolysis- and complement-related proteins (e.g. CFD, F9, CFHR4, FGA).

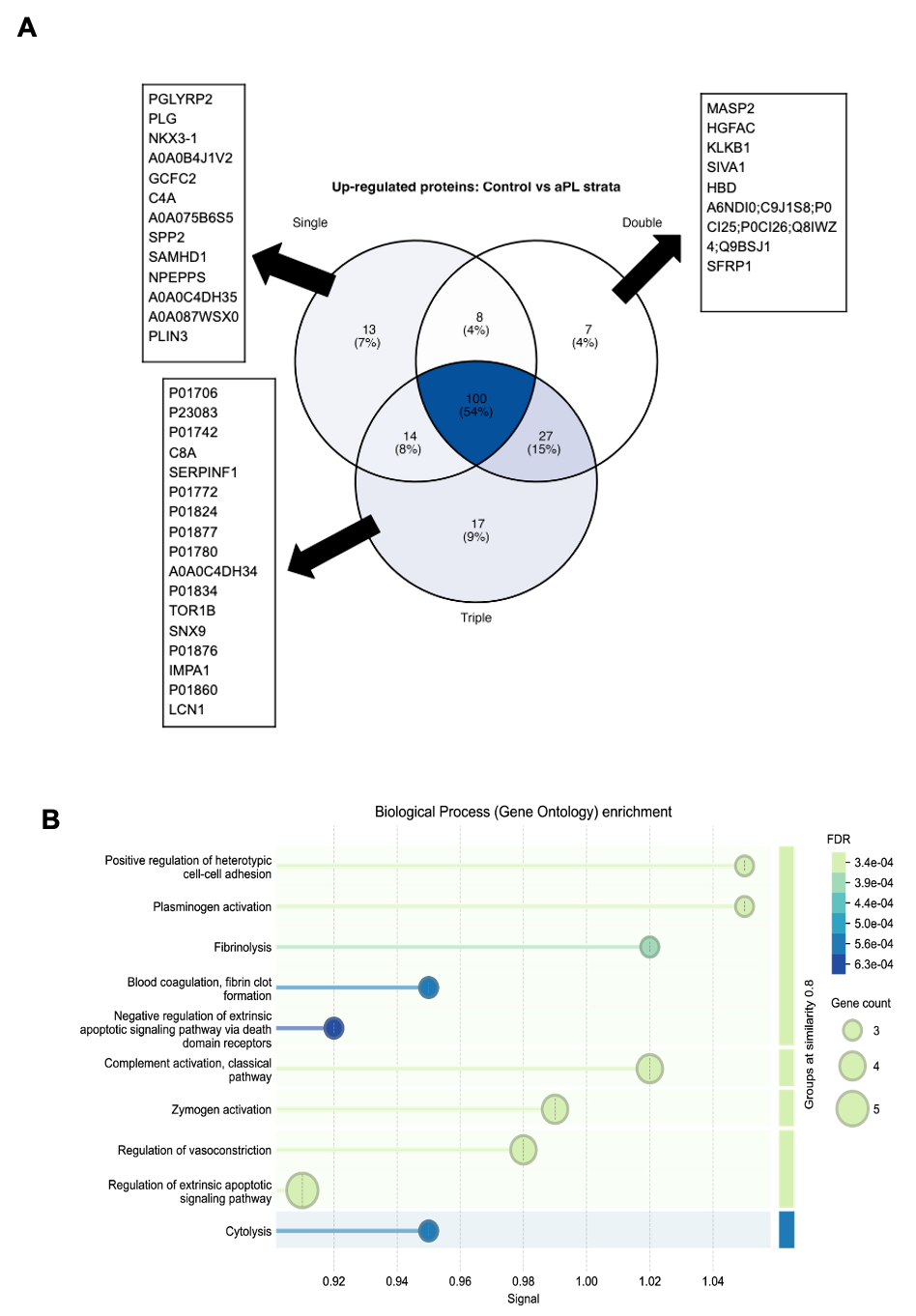

**Supplementary Figure 3.** Proteomic alterations associated with antiphospholipid antibody burden in thrPAPS. (A) Venn diagram showing unique and shared upregulated plasma proteins across single-, double-, and triple-positive aPL profiles compared with healthy controls, with a shared 100-protein core signature and progressive expansion of differentially abundant proteins with increasing aPL burden. (B) Functional enrichment of proteins upregulated in triple-positive APS and of proteins shared between the triple- and double-positive groups, highlighting fibrinolysis, complement activation, and clot formation pathways.

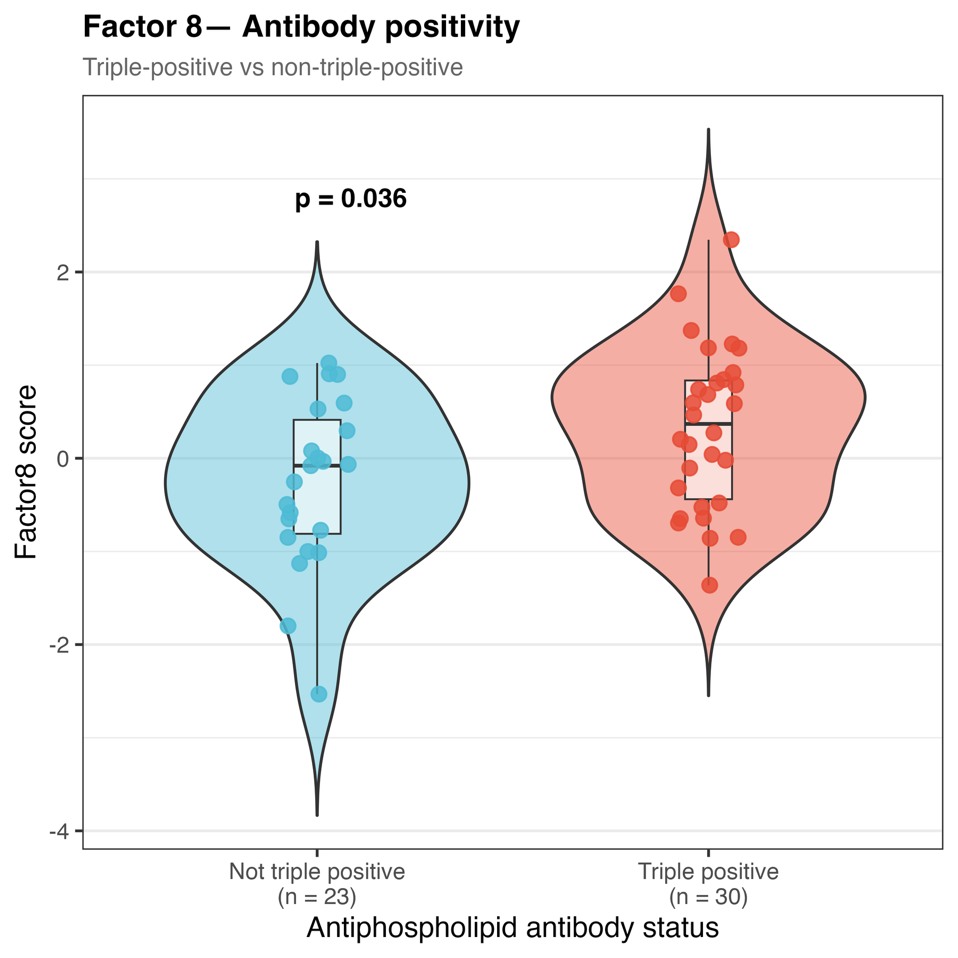

**Supplementary Figure 4:** Factor 8 scores are elevated in triple-aPL-positive patients. Violin and overlaid box plots of MOFA Factor 8 scores in patients stratified by antiphospholipid antibody status (not triple-positive, n = 23; triple-positive, n = 30; two patients without complete aPL typing are excluded, accounting for the difference from the full cohort of n = 55). Boxes show the median and interquartile range; whiskers extend to 1.5× IQR; points represent individual patients. Triple-positive patients had significantly higher Factor 8 scores than non-triple-positive patients (Wilcoxon rank-sum test, p = 0.036), consistent with the monotonic increase of this complement-enriched proteomic programme with aPL burden.
